# Risk Factors for Sudden Cardiac Arrest and Ventricular Arrhythmias in Arrhythmogenic Mitral Valve Prolapse Syndrome

**DOI:** 10.1101/2023.10.06.23296682

**Authors:** Apurba Chakrabarti, John R. Giudicessi, Fatima M. Ezzeddine, Francesca N. Delling, Shalini Dixit, Yoo Jin Lee, Daniele Muser, Silvia Magnani, Aniek Van Wijngaarden, Nina Ajmone Marsan, Marc A. Miller, Jonathan Gandhi, Maria G. Trivieri, Jonaz Font, Raphael Martins, James A. McCaffrey, Pasquale Santangeli, Francis E. Marchlinski, Himal Chapagain, Don Mathew, Krishna Kancharla, Faisal F. Syed, Ahad Abid, Lukasz Cerbin, Wendy S. Tzou, Lohit Garg, Domenico G. Della Rocca, Andrea Natale, Sanghamitra Mohanty, Seth H. Sheldon, Ling Kuo, Kristina H. Haugaa, Eivind W. Aabel, Andres Enriquez, Shingo Maeda, Amrish Deshmukh, Michael Ghannam, Frank M. Bogun, Michael J. Ackerman, Jackson J. Liang

## Abstract

**Background:** Patients with the arrhythmogenic mitral valve prolapse syndrome (AMVPS) are at increased risk for life-threatening ventricular arrhythmias (VAs), but studies have been limited by small sample sizes. We sought to assemble an international AMVPS registry to delineate clinical, imaging, treatment characteristics, and risk factors for sudden cardiac arrest (SCA).

**Methods:** We retrospectively identified two groups of subjects with AMVPS: 1) the MVP-SCA group with SCA, sustained ventricular tachycardia (VT), and ventricular fibrillation (VF); and 2) the MVP-PVC group with significant premature ventricular complexes (PVCs) only. Deidentified data was abstracted locally and combined centrally.

**Results:** We included 217 subjects with AMVPS: 148 (68%) had SCA or VT/VF (MVP-SCA group) and 69 (32%) had PVCs only (MVP-PVC group). Phenotypically, both groups were similar [mean age 44.2±16.7 years, 66% female, 76% with bileaflet prolapse, 55% with mitral annular disjunction (MAD)]. Syncope was more common in the MVP-SCA group than the MVP-PVC group (47% vs 22%, p=0.001) as were anterolateral T-wave inversions (TWIs, 22% vs 7%, p=0.011). Prior mitral valve surgery was less common in the MVP-SCA group (6% vs 20%, p=0.002). These differences remained significant after multivariable adjustment. An electrophysiology (EP) study was negative in 15/45 (33%) of the MVP-SCA subjects.

**Conclusions:** In this international registry, AMVPS subjects were young, female, and had bileaflet prolapse with MAD. A history of syncope and anterolateral TWIs were associated with SCA. Prior mitral valve surgery was less common in SCA subjects. A negative EP study had limited negative predictive value in high-risk patients.

## INTRODUCTION

Mitral valve prolapse (MVP) is a common cause of primary mitral regurgitation worldwide, affecting 1-3% of the general population.^1–3^ While the majority of patients with MVP have benign outcomes, a subset of patients may experience valvular or arrhythmic complications. Valvular complications include endocarditis, thromboembolism, and stroke as well as valvular heart failure and need for mitral valve surgery.^4–6^

Recently, increasing attention has been placed on arrhythmic complications which include sudden cardiac arrest (SCA), ventricular tachycardia (VT), ventricular fibrillation (VF), and symptomatic premature ventricular complexes (PVCs). MVP patients with these arrhythmias and no other plausible alternative etiologies have been labeled with the arrhythmogenic MVP syndrome (AMVPS). Community dwelling patients with MVP experience SCA at rates as low as 0.14 per 100 patient-years.^3^ However, rates may be 0.4% or even higher in those referred to tertiary centers or with risk factors.^4,6^ Given the relatively high prevalence of MVP, even a low incidence of SCA and VT/VF translates into a significant public health concern. Indeed, autopsy studies of younger patients with otherwise unexplained SCA have shown prevalence of MVP between 2-11%.^4,8,9^

Multiple prior studies have sought to identify risk factors to discriminate those at high risk of arrhythmic complications from those with a more benign outcome. These risk factors are electrocardiographic, cardiac monitoring, imaging abnormalities such as inferolateral T-wave inversions (TWIs), non-sustained VT and complex PVCs,^3,7,9,10^ thickened bileaflet prolapse with Barlow’s valve morphology and severe mitral regurgitation,^10–12^ and papillary muscle and left ventricular (LV) delayed enhancement on cardiac magnetic resonance imaging (CMR).^7,13^ Recent research has shown that abnormal separation of the mitral annulus from the basal left ventricular myocardium, termed mitral annular disjunction (MAD), is associated with SCA and VT/VF.^14–18^

To date, most studies of patients with AMVPS have utilized small sample sizes from single centers or populations with a small proportion of patients with SCA. These limitations have made it difficult to characterize the general AMVPS population at highest risk for SCA. We therefore sought to create an international registry of patients with MVP and SCA, VT or VF, or significant PVCs and to define the clinical, imaging, and treatment characteristics as well as risk factors for SCA.

## METHODS

### Patient population

Patients with MVP and VAs were retrospectively identified at 20 international centers. MVP was defined according to the standard definition, with ≥2 millimeters of displacement of the mitral valve leaflets into the left atrium during systole by transthoracic cardiac imaging.^19^ Ventricular arrhythmias (VAs) included unexplained SCA, or documented sustained monomorphic VT, polymorphic VT, VF, and significant PVCs. Significant PVCs were defined as symptomatic PVCs, pleomorphic or bigeminal PVCs, PVCs with a >5% burden, or PVCs requiring anti-arrhythmic drug or catheter ablation therapy. The index enrollment event was the time of SCA or VT/VF in those with sustained VA or SCA. In patients enrolled for significant PVCs, the time of first evaluation for PVCs at the participating institution was considered the enrollment event.

VAs were attributed to MVP after evaluation excluding other known causes. Patients with cardiomyopathy unrelated to MVP, arrhythmic syndromes, channelopathies, and prior myocardial infarction were excluded. The study was approved by the Institutional Review Board of the coordinating center (University of Michigan).

### Data abstraction

De-identified data was collected retrospectively at the individual institutional level into a standardized worksheet and deidentified data were transmitted to a central database for analysis. Data abstracted included baseline demographics, clinical comorbidities and medications, presence of cardiac implantable electronic devices, electrocardiography, echocardiography, ambulatory cardiac monitoring, and CMR. We also examined findings from invasive testing and treatment when performed, including electrophysiology study, catheter ablation procedures, and cardiac surgery. Testing procedures were done according to local institutional protocols and were not standardized across the cohort. Not all subjects had all types of imaging or testing done.

For analysis, inferolateral T-wave inversions (TWI) were defined as T-wave inversion in at least two of leads II, III, aVF, I and aVL. Anterolateral TWI was defined as inversions in leads V5 or V6. A Barlow morphology valve was defined as those with bileaflet prolapse with leaflets at least 5mm in thickness. The grading of mitral regurgitation was determined based on local institutional practice. Mitral annular disjunction (MAD) was defined as measurable systolic separation of the mitral valve leaflet insertion point from the basal left ventricular myocardium with excursion into the left atrium.

The patient population was stratified based on the presence of SCA or sustained VT/VF (MVP- SCA) versus those with significant PVCs without SCA or VT/VF (MVP-PVC).

### Statistical analysis

Continuous variables were reported as mean ± standard deviation and were compared by Student’s t-test. Categorical variables were reported as percentages and compared using chi-square tests. If any analysis group included fewer than 25 patients and failed a Shapiro-Wilks normality test, we utilized non-parametric tests including a Mann-Whitney test for continuous variables and the Fisher’s exact test for categorical variables.

We also performed adjustment with multivariable logistic regression. We included variables that had significant differences (p<0.05) between the MVP-SCA and MVP-PVC groups. We only included variables where the difference could not be explained by inclusion criteria (*e.g.* PVC group including patients with palpitations or higher PVC burden). Sensitivity analyses using logistic regression models using plausibly significant characteristics that were not statistically different were also done. These models included the following variables: age, sex, bileaflet prolapse, MAD, left ventricular ejection fraction by echocardiography, inferolateral TWI as well as prior beta blocker prescription, history of syncope, prior mitral valve surgery, and anterolateral TWI. Given the possibility of interaction between age and mitral valve surgery (*i.e.* the probability of SCA event and mitral valve surgery might be different if the subject required mitral valve surgery at age 30 or age 70 years), models were created with and without mitral valve surgery and age interaction term. Of note, CMR results were not used in multivariable adjustment due to the significantly smaller sample size of patients who completed CMR. Statistical analysis was completed with STATA version 14.2 (College Station, Texas, USA).

## RESULTS

### Demographics

We identified 217 subjects across 20 institutions (Table 1). A total of 148 (68.2%) subjects had prior SCA, sustained VT, or VF and these patients comprised the MVP-SCA group. Specifically, 96 subjects (44.2%) had SCA, 43 subjects (19.8%) had monomorphic VT, 23 subjects (10.6%) had polymorphic VT, 75 subjects (34.6%) had VF, and 8 subjects (3.7%) had electrical storm (Figure 1). Some subjects had more than one VA. Sixty-nine subjects (31.8%) had significant PVCs without SCA or VT/VF, and these patients comprised the MVP-PVC group. Overall, most subjects were young (44.2 ± 16.7 years) and female gender (n=144, 66.4%) with no differences between the groups.

**Figure 1.**
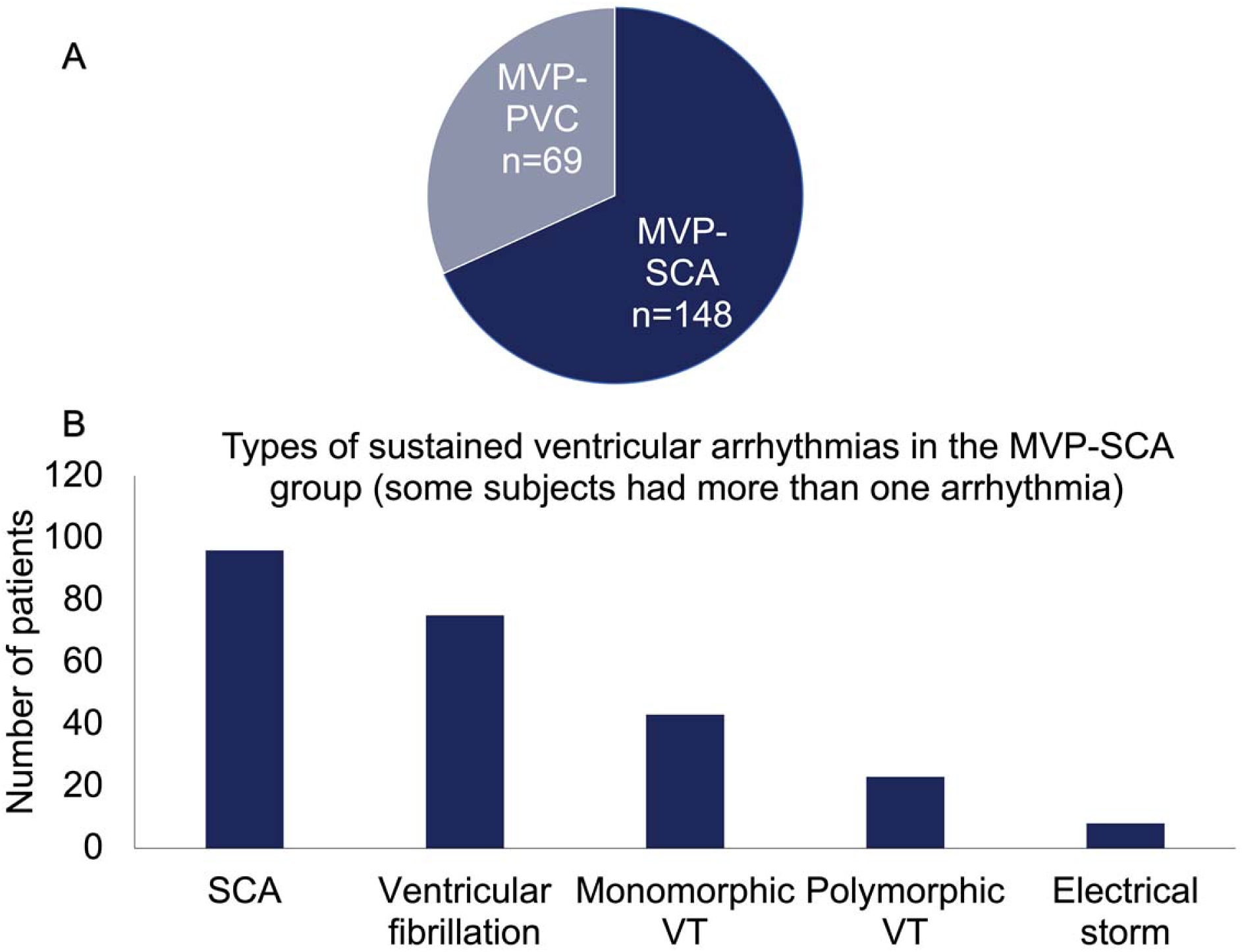
Ventricular arrhythmias in the AMVPS registry. A) The proportions of patients in the MVP-PVC versus MVP-SCA groups. B) Specific ventricular arrhythmias for the MVP-SCA group. Some subjects had more than one type of ventricular arrhythmia.

**Table 1.**
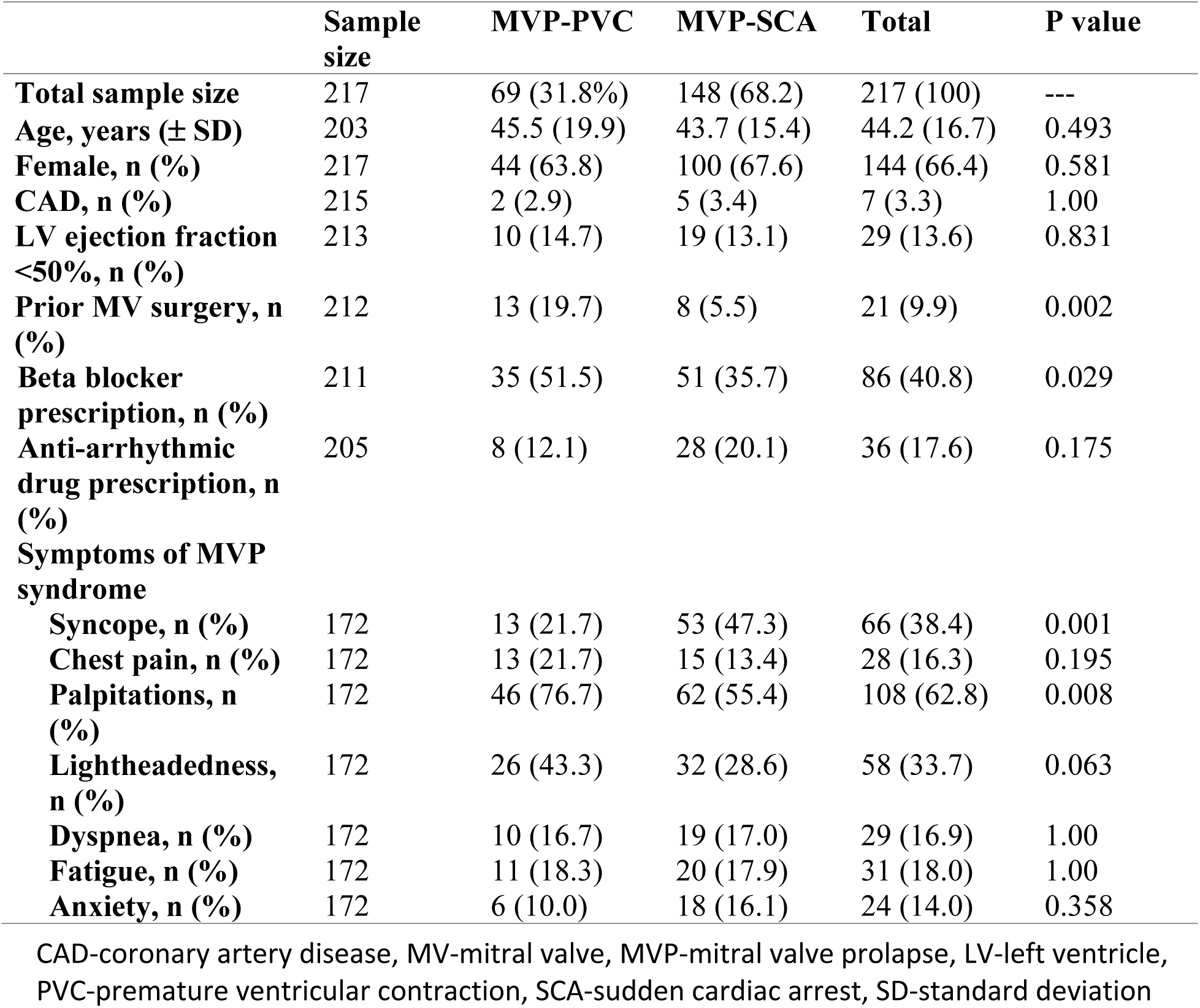
Demographic, clinical, and symptom characteristics.

### Background therapy

Prior mitral valve repair/replacement was significantly less common in the MVP-SCA group (n=8, 5.5%) compared to the MVP-PVC group (n=13, 19.7%, p=0.002, Table 1). Beta blockers were less commonly prescribed in the MVP-SCA group compared to the MVP-PVC group [n=51 (35.7%) versus n=35 (51.5%), respectively, p=0.029]. There was no difference in prescription of anti-arrhythmic drug therapy between MVP-SCA and MVP-PVC groups.

### Mitral valve apparatus

Combined data from echocardiography and cardiac magnetic resonance imaging (CMR) showed that out of 204 patients with data on prolapsing leaflets, bileaflet prolapse was present in most patients in both groups [MVP-SCA n=103 (73.1%), MVP-PVC n=53 (84.1%), p=0.108]. A smaller subset of 152/217 (70.0%) patients had data on mitral valve leaflet thickness and morphology. Of those 152 subjects, 99 (65.1%) had a Barlow morphology with no differences between groups [MVP-SCA n=73 (67.6%), MVP-PVC n=26 (59.1%), p=0.351]. Less than half of patients had moderate or worse mitral valve regurgitation (Table 2). MAD (from either echocardiography or CMR) was present in a little over half patients [MVP-SCA n=82 (55.4%), MVP-PVC n=38 (55.1%), p=0.963, Figure 2B]. There were no statistically significant differences between the MVP-SCA and MVP-PVC groups for mitral valve characteristics.

**Figure 2.**
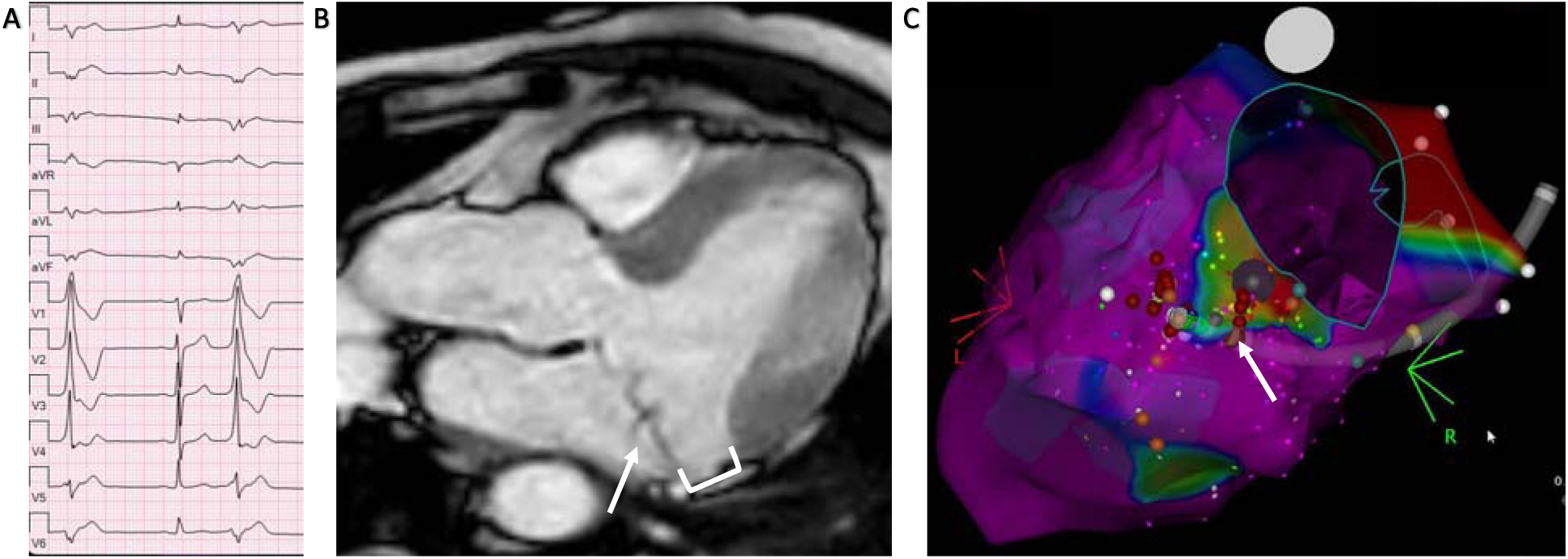
Representative AMVPS subject with PVC bigeminy, bileaflet MVP, MAD and mitral annular origin of PVCs. A) Frequent right bundle superior axis PVCs B) CMR 3 chamber view of the same subject with bileaflet MVP (arrow) and MAD (bracket) C) Electroanatomic map created during invasive catheter ablation showing bipolar voltage and identifying a matching focus (arrow) of the PVC at the mitral annulus in an area of abnormal voltage.

**Table 2.**
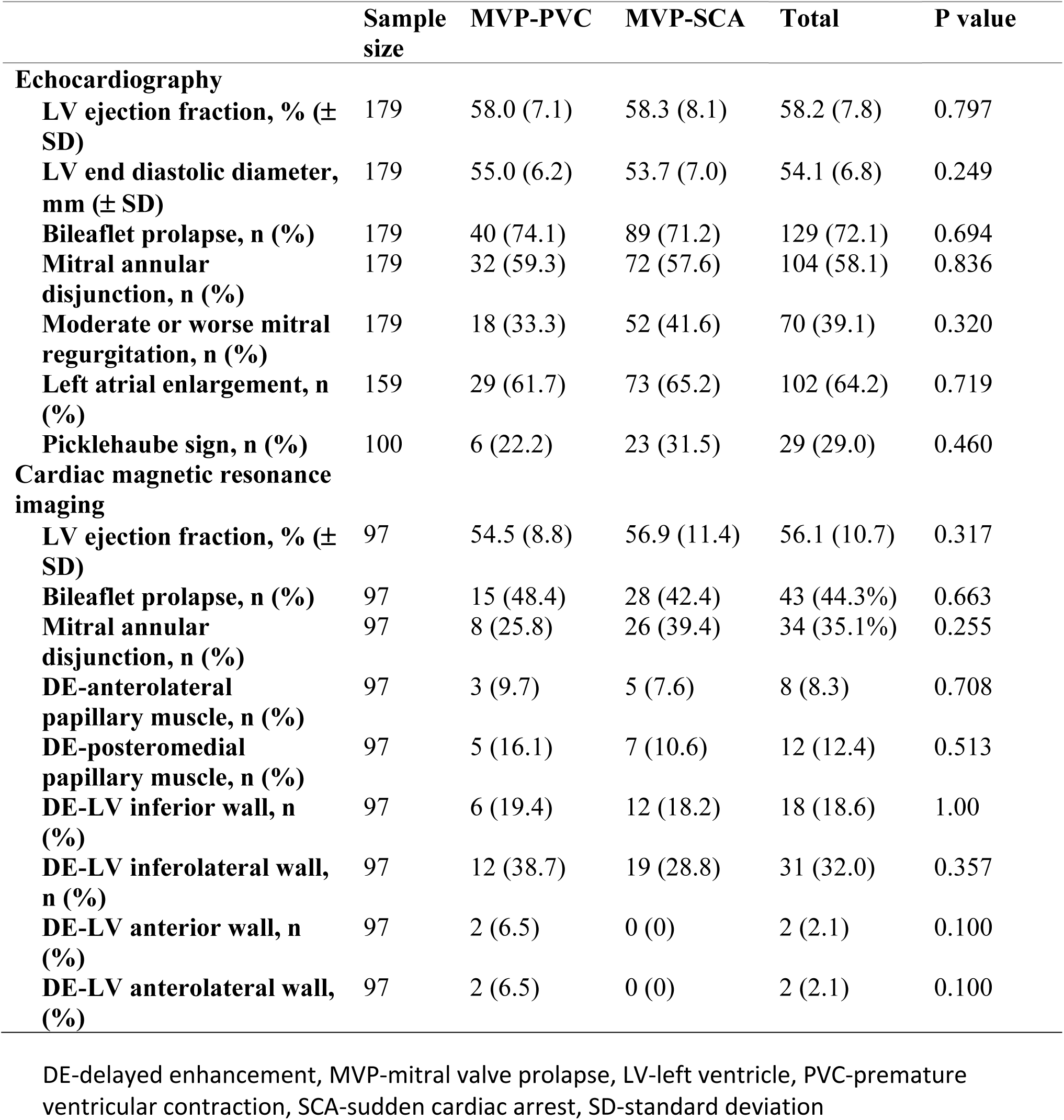
Echocardiographic and cardiac magnetic resonance imaging (CMR) results.

|  | Sample size | MVP-PVC | MVP-SCA | Total | P value |
| --- | --- | --- | --- | --- | --- |
| **Echocardiography** |  |  |  |  |  |
| LV ejection fraction, % (± SD) | 179 | 58.0 (7.1) | 58.3 (8.1) | 58.2 (7.8) | 0.797 |
| LV end diastolic diameter, mm (± SD) | 179 | 55.0 (6.2) | 53.7 (7.0) | 54.1 (6.8) | 0.249 |
| Bileaflet prolapse, n (%) | 179 | 40 (74.1) | 89 (71.2) | 129 (72.1) | 0.694 |
| Mitral annular disjunction, n (%) | 179 | 32 (59.3) | 72 (57.6) | 104 (58.1) | 0.836 |
| Moderate or worse mitral regurgitation, n (%) | 179 | 18 (33.3) | 52 (41.6) | 70 (39.1) | 0.320 |
| Left atrial enlargement, n (%) | 159 | 29 (61.7) | 73 (65.2) | 102 (64.2) | 0.719 |
| Picklehaube sign, n (%) | 100 | 6 (22.2) | 23 (31.5) | 29 (29.0) | 0.460 |
| **Cardiac magnetic resonance imaging** |  |  |  |  |  |
| LV ejection fraction, % (± SD) | 97 | 54.5 (8.8) | 56.9 (11.4) | 56.1 (10.7) | 0.317 |
| Bileaflet prolapse, n (%) | 97 | 15 (48.4) | 28 (42.4) | 43 (44.3%) | 0.663 |
| Mitral annular disjunction, n (%) | 97 | 8 (25.8) | 26 (39.4) | 34 (35.1%) | 0.255 |
| DE-anterolateral papillary muscle, n (%) | 97 | 3 (9.7) | 5 (7.6) | 8 (8.3) | 0.708 |
| DE-posteromedial papillary muscle, n (%) | 97 | 5 (16.1) | 7 (10.6) | 12 (12.4) | 0.513 |
| DE-LV inferior wall, n (%) | 97 | 6 (19.4) | 12 (18.2) | 18 (18.6) | 1.00 |
| DE-LV inferolateral wall, n (%) | 97 | 12 (38.7) | 19 (28.8) | 31 (32.0) | 0.357 |
| DE-LV anterior wall, n (%) | 97 | 2 (6.5) | 0 (0) | 2 (2.1) | 0.100 |
| DE-LV anterolateral wall, n (%) | 97 | 2 (6.5) | 0 (0) | 2 (2.1) | 0.100 |

DE-delayed enhancement, MVP-mitral valve prolapse, LV-left ventricle, PVC-premature ventricular contraction, SCA-sudden cardiac arrest, SD-standard deviation

### Symptoms and the AMVPS

Historical data about the presence of symptoms associated with the AMVPS were available in 172 (79.3%) subjects (Table 1). Syncope was significantly more common in the MVP-SCA versus MVP-PVC group [n=53 (47.3%) versus n=13 (21.7%), p=0.001]. Palpitations were significant less common in the MVP-SCA group (n=62, 55.4%) compared to the MVP-PVC group (n=46, 76.7%, p=0.008). Lightheadedness was numerically less common in the MVP-SCA versus MVP-PVC group [n=32 (28.6%) versus n=26 (43.3%), respectively] but the difference was non-significant with a p=0.063. There were no differences in the prevalence of symptoms of chest pain, near syncope, dyspnea, fatigue, or anxiety.

### Electrocardiography

In the 174 subjects (80.1%) where electrocardiographic (ECG) data was available, the corrected QT interval in the MVP-SCA group was 438 ± 24 msec versus 445 ± 25 msec in the MVP-PVC group (p=0.072, Table 3). While present in over half of patients, there was no difference in the presence of inferolateral TWIs [MVP-SCA n=69 (59.5%), MVP-PVC n=29 (50.0%), p=0.234]. While anterolateral TWIs were less frequently seen in the overall cohort (n=30, 17.2%), they were present more often in the MVP-SCA group (n=26, 22.4%) than the MVP-PVC group (n=4, 6.9%, p=0.011).

**Table 3.**
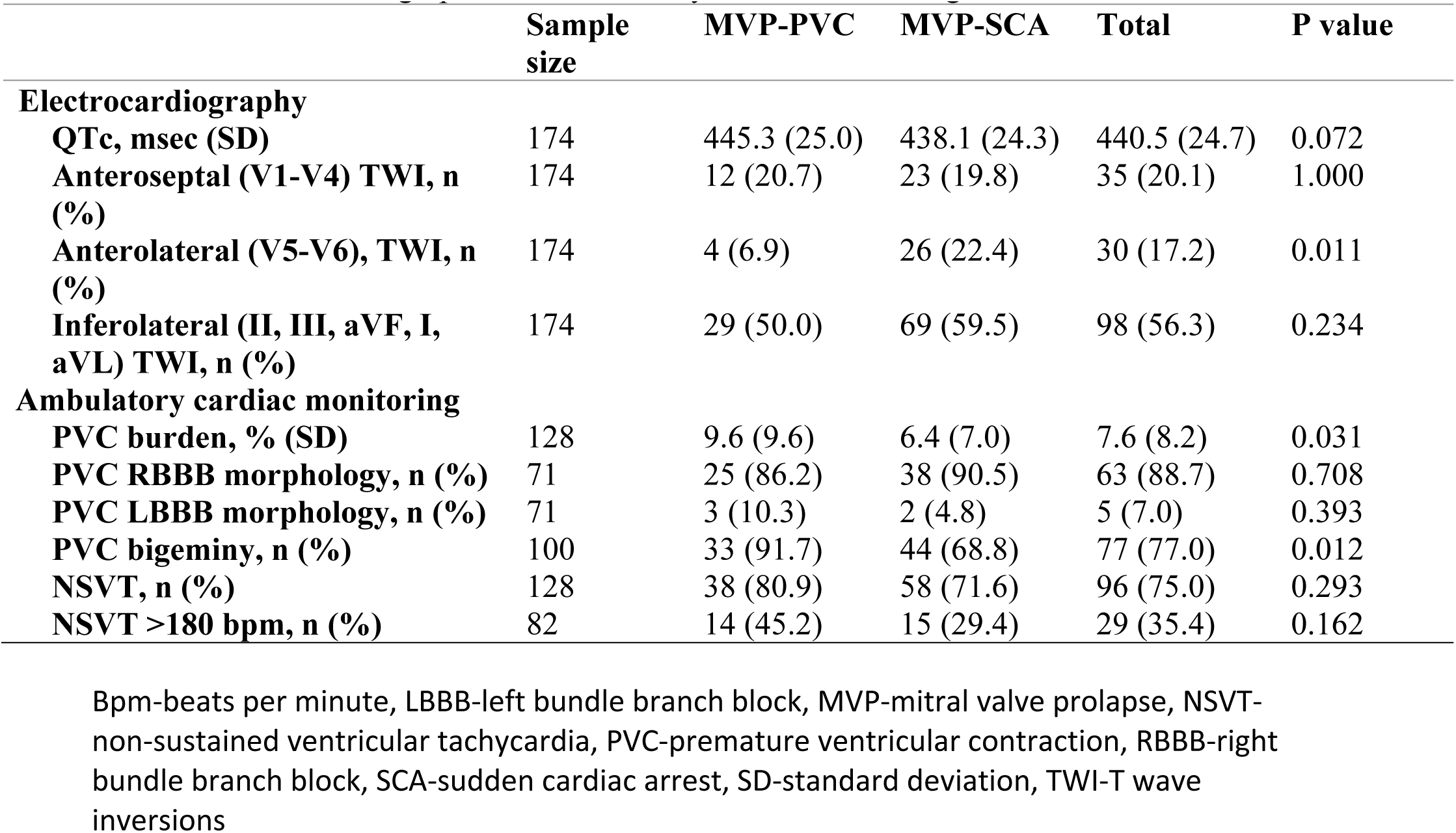
Electrocardiographic and ambulatory cardiac monitoring results.

|  | **Sample**<br>**size** | **MVP-PVC** | **MVP-SCA** | **Total** | **P value** |
| --- | --- | --- | --- | --- | --- |
| **Electrocardiography** |  |  |  |  |  |
| **QTc, msec (SD)** | 174 | 445.3 (25.0) | 438.1 (24.3) | 440.5 (24.7) | 0.072 |
| **Anteroseptal (V1-V4) TWI, n (%)** | 174 | 12 (20.7) | 23 (19.8) | 35 (20.1) | 1.000 |
| **Anterolateral (V5-V6), TWI, n (%)** | 174 | 4 (6.9) | 26 (22.4) | 30 (17.2) | 0.011 |
| **Inferolateral (II, III, aVF, I, aVL) TWI, n (%)** | 174 | 29 (50.0) | 69 (59.5) | 98 (56.3) | 0.234 |
| **Ambulatory cardiac monitoring** |  |  |  |  |  |
| **PVC burden, % (SD)** | 128 | 9.6 (9.6) | 6.4 (7.0) | 7.6 (8.2) | 0.031 |
| **PVC RBBB morphology, n (%)** | 71 | 25 (86.2) | 38 (90.5) | 63 (88.7) | 0.708 |
| **PVC LBBB morphology, n (%)** | 71 | 3 (10.3) | 2 (4.8) | 5 (7.0) | 0.393 |
| **PVC bigeminy, n (%)** | 100 | 33 (91.7) | 44 (68.8) | 77 (77.0) | 0.012 |
| **NSVT, n (%)** | 128 | 38 (80.9) | 58 (71.6) | 96 (75.0) | 0.293 |
| **NSVT >180 bpm, n (%)** | 82 | 14 (45.2) | 15 (29.4) | 29 (35.4) | 0.162 |

Bpm-beats per minute, LBBB-left bundle branch block, MVP-mitral valve prolapse, NSVT-non-sustained ventricular tachycardia, PVC-premature ventricular contraction, RBBB-right bundle branch block, SCA-sudden cardiac arrest, SD-standard deviation, TWI-T wave inversions

### Ambulatory cardiac monitoring

One hundred twenty-eight subjects (59.0%) had data from ambulatory cardiac monitoring (Table 3). The PVC burden was significantly lower in the MVP-SCA group versus the MVP-PVC group (6.4% ± 7.0% vs 9.6% ± 9.6%, p=0.031). Most subjects had PVCs with a right bundle branch (RBBB, Figure 2A) morphology (n=63, 88.7%) with no difference between groups. Bigeminal PVCs were significantly less common in the MVP-SCA group. Most patients had at least one episode of one non-sustained VT (n=96, 75.0%), but there was no difference between groups. Eighty-two of 128 subjects had data on the maximum rate of episodes of non-sustained VT. There was no difference in the prevalence of patients with a maximum rate of non-sustained VT>180 beats per minute (bpm) between groups.

### Echocardiography

One hundred seventy-nine subjects (82.5%) had complete data from echocardiography available (Table 2). Overall, subjects in both groups had normal LV ejection fraction and LV size with no differences between groups. Most patients did have left atrial enlargement but there was no difference between groups. One-hundred patients (46.1%) had data on the presence of Picklehaube sign.^20,21^ Twenty-nine patients (29.0%) had the Picklehuabe sign, but there was no difference between groups.

### Cardiac magnetic resonance imaging (CMR)

Ninety-seven subjects (44.7%) had contrast enhanced CMR data available (Table 2). There were no differences in delayed enhancement (DE) distribution between the MVP-SCA and MVP-PVC groups. Thirteen subjects (13.4%) had DE involving either the anterolateral or posteromedial papillary muscles. More subjects had DE of either inferior (n=18, 18.6%) or inferolateral (n=31, 32.0%) LV myocardium. Forty-five subjects (46.4%) had at least one area of DE. In our cohort, 99 subjects (45.6%) had information on presence of inferior or inferolateral DE on CMR and TWI on surface ECG. A total of 40/99 (40.4%) patients had DE in the inferior or inferolateral LV myocardium. Of these 31/40 (77.5%) patients had inferolateral TWI and only 9/40 (22.5%) had DE but no corresponding inferolateral TWI (p=0.019). Of the 2/97 (4.1%) that had anterolateral DE in the LV myocardium, none had associated anterolateral TWI.

### Electrophysiology study

A total of 150 subjects underwent an electrophysiology study (EPS) with programmed ventricular stimulation. Subjects in the MVP-SCA group were less likely to undergo EPS than in the MVP-PVC group (Table 4). In those that did undergo an EPS, there was no difference in the prevalence of inducible VT/VF between groups. Importantly, of the 45 subjects in the MVP- SCA group who had prior SCA or sustained VT/VF and underwent EPS, 15/45 (33.3%) were non-inducible during an EPS despite a prior arrhythmic event. Of those in the MVP-SCA group with a negative EPS, 6/15 had prior monomorphic VT, 1/15 had polymorphic VT, 7/15 had VF and one had an unknown arrhythmia inciting SCA.

**Table 4.** Percutaneous catheter ablation, cardiac implantable device therapy, and surgical treatments.

|  | **Sample size** | **MVP-PVC** | **MVP-SCA** | **Total** | **P value** |
| --- | --- | --- | --- | --- | --- |
| **ICD placed during follow-up, n (%)** | 216 | 33 (47.8) | 128 (87.1) | 161 (74.5) | <0.001 |
| **MV surgery during follow-up, n (%)** | 195 | 3 (5.6) | 11 (7.8) | 14 (7.2) | 0.761 |
| **EPS performed, n (%)** | 150 | 27 (64.3) | 45 (41.7) | 72 (48.0) | 0.018 |
| **EPS positive, n (%)** | 72 | 12 (44.4) | 30 (66.7) | 42 (58.3) | 0.085 |
| **PVC ablation, n (%)** | 207 | 35 (54.7) | 50 (35.0) | 85 (41.1) | 0.008 |
| **VT ablation, n (%)** | 146 | 8 (18.2) | 18 (17.7) | 26 (17.8) | 1.000 |

EPS-electrophysiology study, ICD-implantable cardiac defibrillator, MV-mitral valve, MVP-mitral valve prolapse, PVC-premature ventricular contraction, SCA-sudden cardiac arrest, VT-ventricular tachycardia

### Mitral valve surgery and device therapy

After the index event, mitral valve repair/replacement was performed in a minority of patients with no differences between either group [MVP-SCA n=11 (7.8%), MVP-PVC n=3, (5.6%), p=0.761, Table 4]. Implantable cardiac defibrillators were placed commonly in both groups after the index event but significantly more in the MVP-SCA group (n=128, 87.1%) versus the MVP- PVC group (n=33, 47.8%, p<0.001).

### Invasive catheter ablation

PVC ablation was performed less often in the MVP-SCA group (n=50, 35.0%) versus the MVP- PVC group (n=35, 54.7%, p=0.008, Table 4). Of the 85 subjects who underwent PVC ablation, the most common sites of origin for PVCs (Figure 3A) were the posteromedial papillary muscle (n=48), anterolateral papillary muscle (n=37), mitral annulus (n=11, Figure 2C), left ventricular (LV) inferolateral wall (n=9), fascicles (n=7), LV septum (n=7), LV inferior wall (n=6), LV outflow tract (n=5), right ventricular (RV) outflow tract (n=5), or LV anterolateral wall (n=2). VT ablation was performed in a minority of patients (Table 4) with no differences between groups. Sites of origin for VTs (Figure 3B) included posteromedial papillary muscle (n=6), fascicles (n=6), mitral annulus (n=6), anterolateral papillary muscle (n=5), LV inferolateral wall (n=4), LV inferior wall (n=3), right ventricular source (n=2), LV apex (n=1), and LV anterolateral wall (n=2). Each subject may have had more than one focus of PVC or VTs.

**Figure 3.**
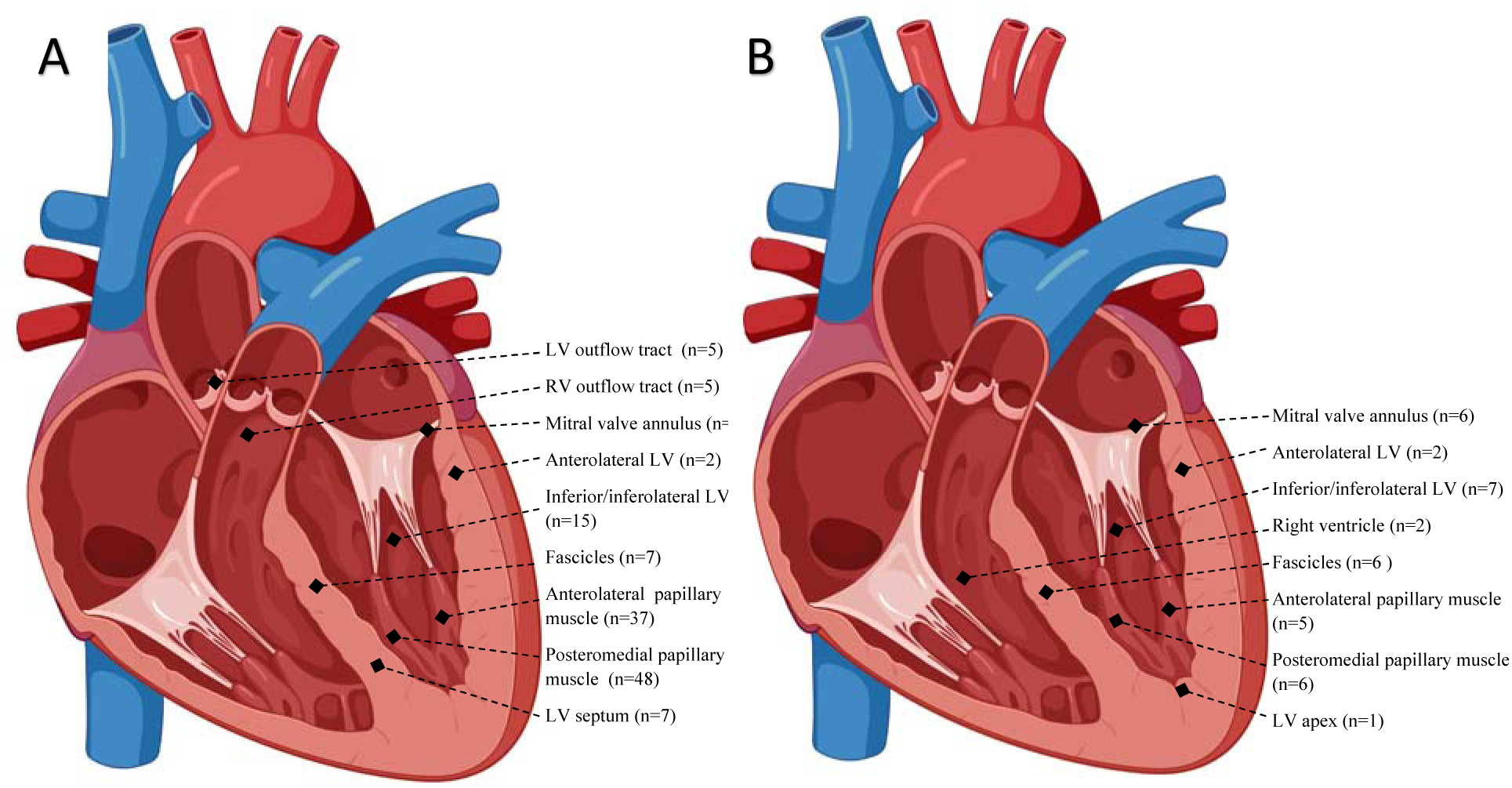
Sites of origin for PVCs and VTs in subjects undergoing catheter ablation. A) Schematic demonstrating sites of origin for PVCs in 85 patients that underwent PVC ablation. B) Sites of origin for VTs in 26 patients that underwent VT ablation. Some subjects had more than one focus of PVC or VT.

### Multivariable risk adjustment

The presence of anterolateral TWIs, a history of syncope, a prior beta blocker prescription, and prior mitral valve surgery had statistically significant (p<0.05) differences between MVP-PVC and MVP-SCA groups. Other variables, such as PVC burden, the presence the PVC bigeminy, and a history of palpitations were also significantly different. However, these differences could be explained by the inclusion criteria for PVC group which included PVC burden >5%, PVC bigeminy, and symptomatic PVCs. Therefore, these variables were excluded. CMR data was also excluded due to smaller sample sizes. A multivariable logistic regression model of the four significant variables showed persistent significance of prior mitral valve surgery [Odds Ratio (OR) 0.17, 95% confidence interval (CI, 0.05, 0.57), p=0.004], anterolateral TWI [OR 5.4, 95% CI (1.6, 17.7), p=0.005], and a history of syncope [OR 3.0, 95% CI (1.4, 6.6), p=0.007] but beta blocker prescription was no longer significant [OR 0.51, 95% CI (0.25, 1.0), p=0.06]. This model is shown in Figure 4.

**Figure 4.**
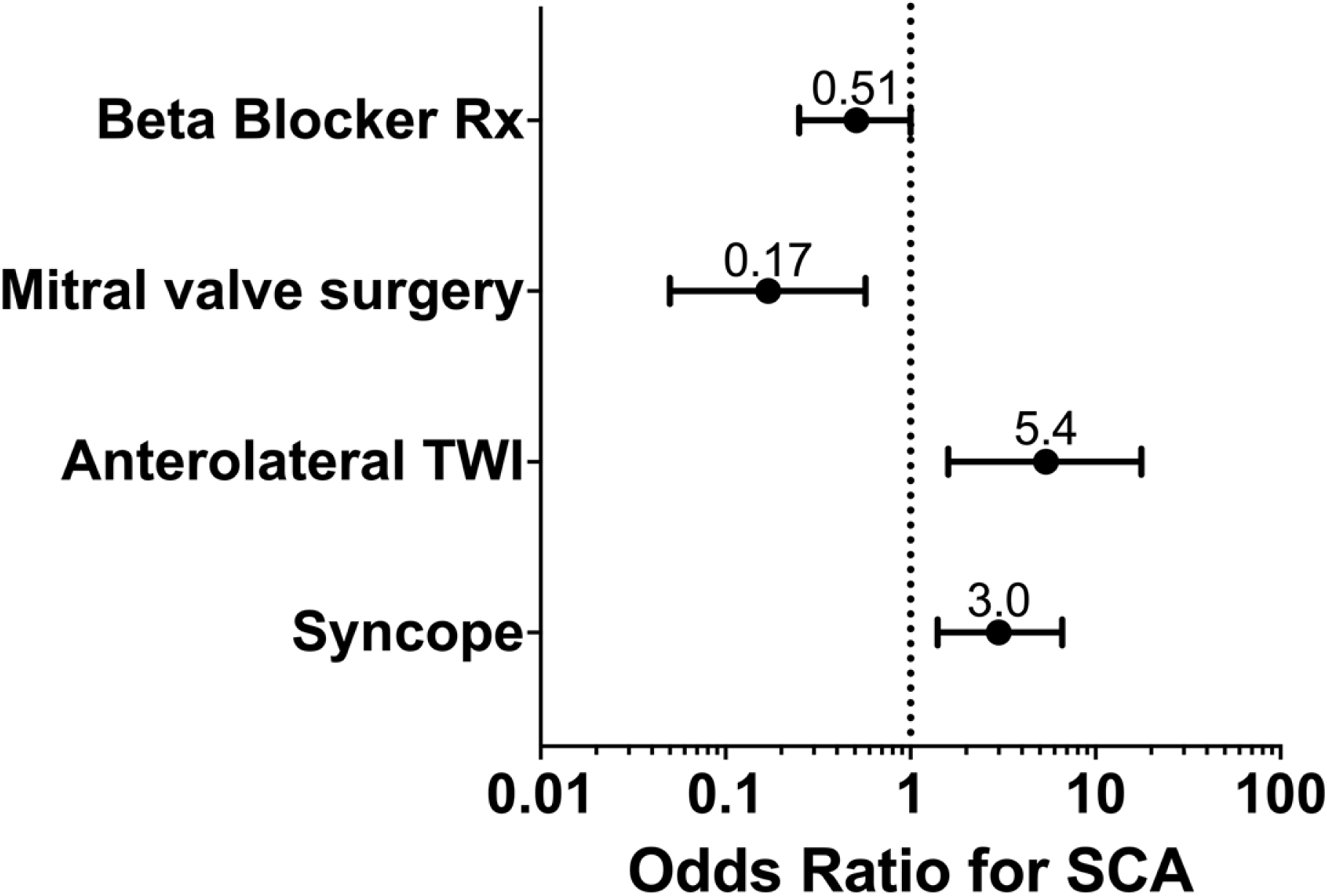
Multivariable logistic regression model. Model analyzing the association of a history of syncope, beta blocker prescription, mitral valve (MV) surgery, and anterolateral TWI with SCA or VT/VF.

Sensitivity analysis was done with additional logistic regression models including other clinically significant variables that were not statistically different between groups (age, sex, bileaflet prolapse, MAD, LV ejection fraction by echocardiography, inferolateral TWI as well as prior beta blocker prescription, history of syncope, prior mitral valve surgery, and anterolateral TWI). These are shown in supplemental data. These combined models showed that the presence of anterolateral TWIs and a history of syncope remained significantly associated with increased SCA and beta blocker prescription remained non-significantly associated with SCA. The association of mitral valve surgery and SCA was significant if its interaction with age was included (see supplemental Tables S1 and S2). No other variables were significantly associated with SCA.

## DISCUSSION

This study represents the largest multicenter series characterizing patients with AMVPS at highest arrhythmic risk, including those with SCA. This cohort of 217 subjects included 96 patients (44%) with SCA and 52 (24%) patients with sustained VT/VF. This study expands upon the initial comprehensive description of AMVPS that stemmed from an analysis of 10/24 SCA survivors with structurally normal hearts that had bileaflet MVP where the key components of AMVPS were defined.^9^ In contrast, the largest previous case series examining AMVPS included 295 patients with some VAs but only 10 patients (3.3%) were SCA survivors.^10^ Although our analysis compared MVP subjects with SCA or VT/VF versus those with significant PVCs, both groups included subjects that would meet multiple risk criteria for SCA (*e.g.* non-sustained VT, bigeminal PVCs, MAD).

In general, our cohort phenotypically characterizes patients with AMVPS as typically younger, female gender, with bileaflet prolapse and MAD (Figure 5). Over half had mild or less mitral regurgitation. Cardiac monitoring shows frequent inferolateral TWIs, frequent PVCs (average 7.6%), typically with RBBB morphology, often with PVC bigeminy and non-sustained VT. Forty-six percent of patients will have some DE on CMR, but only a minority will have papillary muscle LGE in our study. These characteristics have also been previously documented in prior studies.^3,7,9,11,12,15,16,22,23^

**Figure 5.**
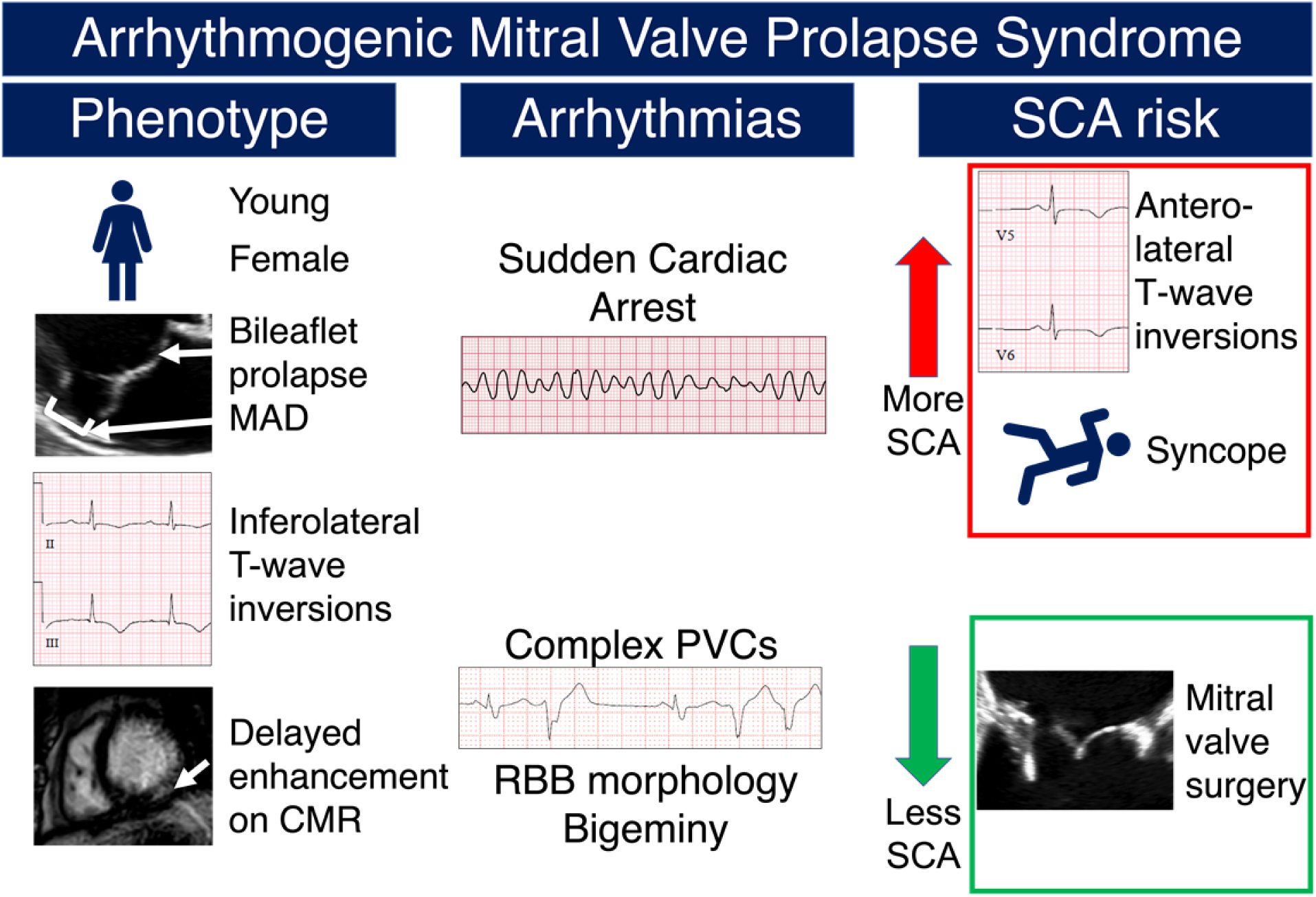
The Arrhythmogenic Mitral Valve Prolapse Syndrome (AMVPS): phenotype, arrhythmias, and risk factors for SCA. Subjects with AMVPS have a typical phenotype with associated clinical, electrocardiographic, echocardiographic findings, and cardiac magnetic resonance imaging (MRI) characteristics (left column). These patients are at risk for otherwise unexplained ventricular arrhythmias including SCA, VT/VF, and significant PVCs (middle column). Clinical and electrocardiographic features more common in MVP patients with SCA or sustained VT/VF compared to PVC patients are shown in the red box (right column, top half) and features less common in SCA patients compared to PVC patients are shown in the green box (right column, bottom half).

Our subjects were predominately female and younger age, which has been seen in most studies enriched with MVP patients with SCA.^7,9,23–25^ In other studies with a lower proportion of patients with SCA, the average age was higher and a greater proportion of males were seen.^16,26^ It is possible that the population at highest risk of SCA is younger and female although not all autopsy studies have shown a female or younger age predominance.^8^

Thus far, there is limited evidence supporting the therapeutic use of any medications for AMVPS. In our study, beta blocker prescription was significantly less common in patients with MVP-SCA than in those with MVP-PVC group. Beta blockers have been recommended in the past for patients with symptomatic VAs, including for patients with MVP,^27^ but to our knowledge there has not been any data supporting their efficacy specifically in MVP patients. The causality of this association remains unclear; it is possible that those in the MVP-PVC group were more frequently treated with beta-blockers for symptomatic PVCs. Indeed, after multivariable adjustment, beta blocker prescription was not associated with reduced chance of SCA. Nevertheless, the possible protective association between beta blockers and SCA or sustained VT/VF warrants further exploration in prospective studies. A small case series has examined the efficacy of flecainide in patients with AMVPS and arrhythmias not responding to beta blockers,^28^ however data on specific anti-arrhythmic drugs was unavailable in our registry.

Studies examining the protective role of mitral valve surgery in treating AMVPS have shown mixed results in the past.^29–33^ In our study, a history of prior mitral valve surgery was significantly less common in MVP-SCA patients although this relationship was weakened after controlling for age (Figure 5). Another large retrospective study of MVP patients showed that the association between VAs and excess mortality was abrogated in MVP patients that underwent mitral valve surgery but not those undergoing medical management.^10^ Others have hypothesized that repeated mechanical strain from the abnormal mitral valve apparatus might contribute to cardiac fibrosis and arrhythmogenesis over time.^34^ It follows logically that mitral valve repair or replacement, which would potentially improve valve mechanics, may limit progression of fibrosis and therefore reduce the onset of serious ventricular arrhythmias. However, it is also possible that this association is from the bias that older survivors are more likely to undergo valve surgery. Given that the majority of the patients in the current series had only mild or less mitral regurgitation, surgical intervention would usually not be indicated for valvular reasons. In patients with valvular indications for surgery, arrhythmic benefits may play a supportive role for surgery, but this concept requires further evaluation in prospective studies.

The existence of a symptom complex associated with MVP is controversial. In our study, a history of syncope was more common in the MVP-SCA than the MVP-PVC group while symptoms such as palpitations and lightheadedness, while somewhat common, were not associated with SCA. This suggests that particular attention should be placed on MVP patients with syncope compatible with an arrhythmic etiology. These patients may warrant further testing. This is consistent with the published expert consensus statement.^35^

Several studies have shown inferolateral TWIs associated with AMVPS and in our population over half of patients did have inferolateral TWIs.^7,16,18^ A novel finding in the current study was that anterolateral TWIs (involving precordial leads V5 or V6), while significantly less prevalent overall, were significantly more common among MVP-SCA subjects compared with MVP-PVC subjects (Figure 5). Anterolateral TWI were not associated with DE on CMR. In contrast, the presence of DE in the LV inferior or inferolateral myocardium was associated with presence inferolateral TWIs. This lends credence to the hypothesis that cardiac fibrosis leads to repolarization abnormalities in the corresponding myocardium and may contribute to electrical instability and arrhythmogenesis.

Electrophysiology studies are performed in a wide variety of patient groups to help stratify risk of VAs. The predictive validity of EPS in patients diagnosed with AMVPS is not well documented. One-third of patients in the current series with a history of SCA or sustained VT/VF still had a negative EPS, suggesting that EPS alone as a diagnostic study may have limited negative predictive value in otherwise high-risk patients. Importantly, most centers perform programmed ventricular stimulation from one or two right ventricular sites. However, given the possibility of site-specificity for programmed ventricular stimulation, since the arrhythmia substrates in MVP-SCA are predominantly in the LV, programmed stimulation from both the right and left ventricles may increase predictive value of EPS.

Our study has several important limitations. First, due to its retrospective nature, causality cannot be inferred. Due to the multicenter design, there was variable work-up performed and access to records; many subjects did not have all forms of testing. Missing data could introduce bias. Additionally, the performance and interpretation of testing, imaging, and treatment could not be standardized. Since our series did not have control group of low or average arrhythmia risk MVP patients, we are unable to specifically identify risk factors that delineate patients at low and high risk of arrhythmias.

## CONCLUSIONS

In this large international registry, subjects with arrhythmogenic mitral valve prolapse syndrome were younger, female, and typically had bileaflet MVP, mitral annular disjunction, inferolateral T-wave inversions, non-sustained VT, and frequent PVCs often with PVC bigeminy. Less than half of patients had abnormal delayed enhancement on cardiac magnetic resonance imaging. A history of syncope and anterolateral T-wave inversions were more common in patients who survived SCA or sustained VT/VF than in those with PVCs alone. A history of prior mitral valve surgery was less common in SCA and VT/VF patients than in PVC patients. A negative EP study had limited negative predictive value in high-risk patients.

## Data Availability

All data available on request

## Abbreviations

AMVPS: arrhythmogenic mitral valve prolapse syndrome
CI: confidence interval
CMR: cardiac magnetic resonance imaging
DE: delayed enhancement
EF: ejection fraction
EPS: electrophysiology study
ICD: implantable cardiac defibrillator
LV: left ventricle
MAD: mitral annular disjunction
MV: mitral valve
MVP: mitral valve prolapse
OR: odds ratio
PVC: premature ventricular contraction
RV: right ventricle
SCA: sudden cardiac arrest
SD: standard deviation
TWI: T wave inversion
VA: ventricular arrhythmias
VF: ventricular fibrillation
VT: ventricular tachycardia

